# Comparison of Bone Augmentation Techniques in the Mandibular Posterior Region: A Randomized Clinical Trial Protocol

**DOI:** 10.1101/2025.11.09.25339853

**Authors:** Antonio Lanata-Flores, Marcelo Armijos Briones, Pablo Benítez Sellán, Patricia Estefanía Ayala Aguirre

## Abstract

This randomized, double-blind, parallel-group clinical trial will compare the effectiveness of three bone-augmentation techniques applied to the posterior mandible using the cortical shell approach. Seventy-eight participants will be randomly allocated into three groups (autogenous, xenogeneic, or allogeneic cortical shells), each combined with a 1:1 mixture of autogenous bone chips and biomaterial.

The study was approved by the CEISH-ITSUP Ethics Committee (Approval No. 1715824376) and is registered at ClinicalTrials.gov (Identifier: NCT06661447). Recruitment has not yet begun.

The primary outcome will be horizontal bone gain (mm) evaluated by cone-beam computed tomography (CBCT) at 3, 6, and 12 months. Volumetric and morphometric analyses will be performed through a standardized digital workflow integrating intraoral scanning, CBCT segmentation, and surface registration. Secondary outcomes will include bone density, surgical duration, and postoperative inflammation.

Based on previous CBCT-based randomized clinical trials, a total of 78 participants (26 per group) will provide 80–90% power to detect a 1.0 mm difference in horizontal bone gain among groups. This study aims to generate controlled, quantitative evidence on the volumetric and clinical performance of different cortical graft materials for posterior mandibular reconstruction.

## INTRODUCTION

Alveolar bone loss following tooth extraction remains one of the main challenges in oral and maxillofacial rehabilitation. Ridge resorption occurs rapidly, particularly at the buccal aspect and compromises prosthetic and esthetic outcomes(1). Several augmentation techniques have been described, including guided bone regeneration (GBR), particulate onlay grafts, autogenous block grafts, ridge splitting, sinus-floor elevation, and distraction osteogenesis, each with distinct advantages and drawbacks related to complexity, healing, and stability (2,3).

Among these, the cortical shell technique provides predictable results for horizontal and three-dimensional reconstruction. Thin cortical plates, harvested mainly from the mandibular ramus or symphysis, are fixed to the recipient ridge with microscrews to form a biologic box that contains particulate graft material (4). This approach combines osteogenic autogenous chips with a xenogeneic scaffold to ensure mechanical stability and volume maintenance (5,6).

However, harvesting autogenous bone can cause donor-site morbidity such as paresthesia, swelling, and limited bone availability (7). To address these issues, xenogeneic and allogeneic cortical laminae have been introduced as ready-to-use alternatives that replicate the mechanical properties of autogenous bone while avoiding secondary surgery. Xenogeneic grafts exhibit favorable biocompatibility and osteoconductivity but show variable remodeling rates(8); allogeneic bone grafts offer biologic compatibility but show variable resorption (9).

Evidence remains limited, as most reports are retrospective and rely on two-dimensional imaging. Recent advances in digital volumetric imaging have made it possible to quantify bone remodeling with high precision. Three-dimensional analysis using cone-beam computed tomography (CBCT) combined with specialized software for segmentation and surface registration allows accurate evaluation of volumetric and morphologic changes in augmented areas. This digital workflow enables standardized measurements over time and provides reproducible data for comparing the performance of different grafting materials. Accordingly, this trial will compare the clinical, radiographic, and volumetric performance of autogenous, xenogeneic, and allogeneic cortical shells in a randomized, double-blind, split-mouth design. By combining digital imaging software for standardized CBCT evaluation with three-dimensional analysis tools for quantitative comparison, the study aims to determine whether prefabricated laminae can achieve outcomes comparable to autogenous plates while reducing surgical morbidity and operative time.

## MATERIALS AND METHODS

### Study design

A randomized, double-blind, parallel-group clinical trial will be conducted at the Universidad de Especialidades Espíritu Santo (UEES), Samborondón, Ecuador, in accordance with the SPIRIT 2025. Ethical approval was granted by CEISH-ITSUP (Approval No. 1715824376). The study has not yet begun participant recruitment, data collection, or surgical procedures. Recruitment and data collection are expected to begin in March 2026 and conclude in December 2026. Participants will be randomly assigned to one of three groups receiving autogenous, xenogeneic, or allogeneic cortical shells. This parallel allocation avoids potential cross-interference between grafts and allows independent comparison among materials. The trial schedule of enrolment, interventions, and assessments is presented in Figure 1, following the SPIRIT 2025 recommendations.

**Figure 1.**
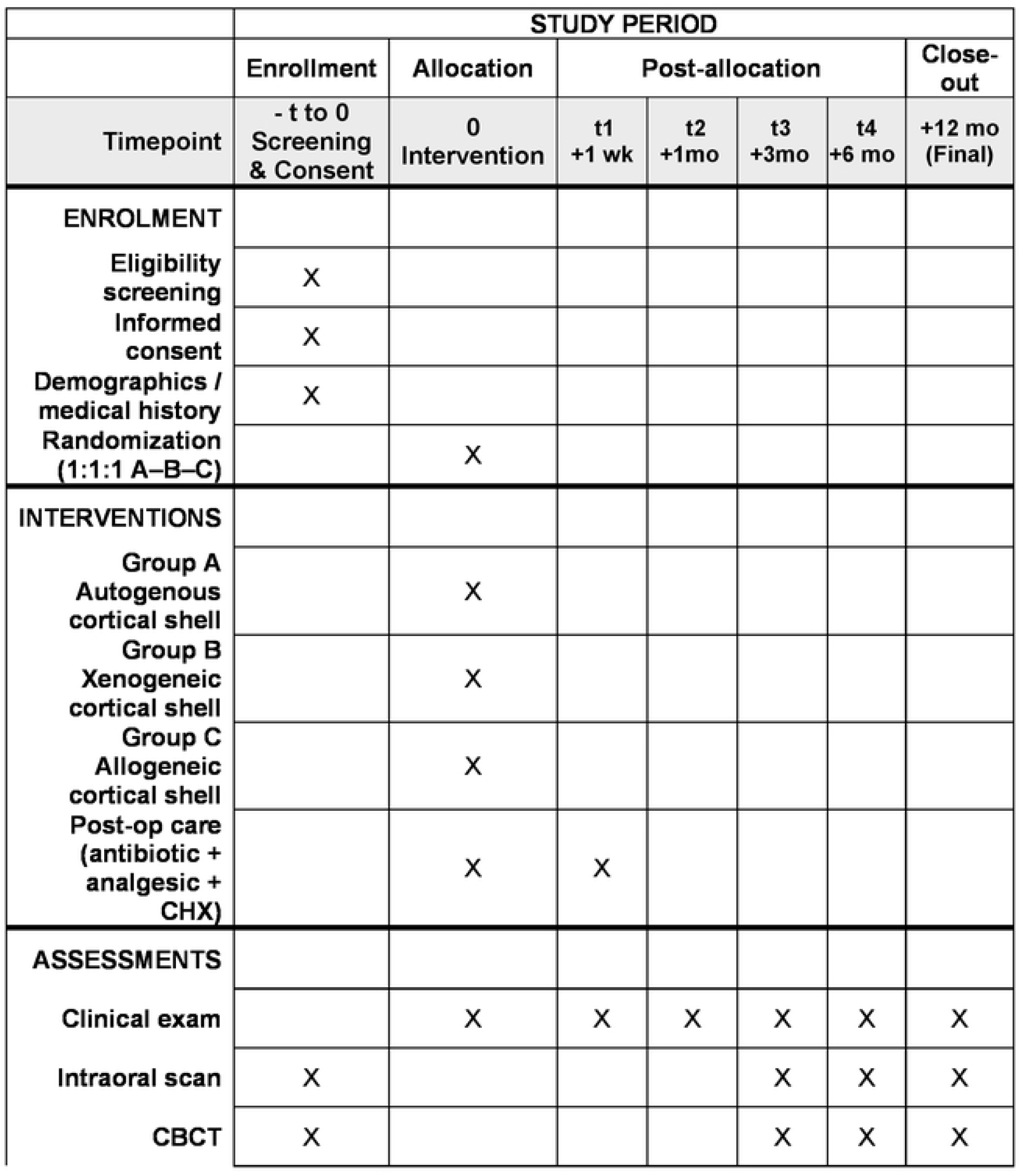
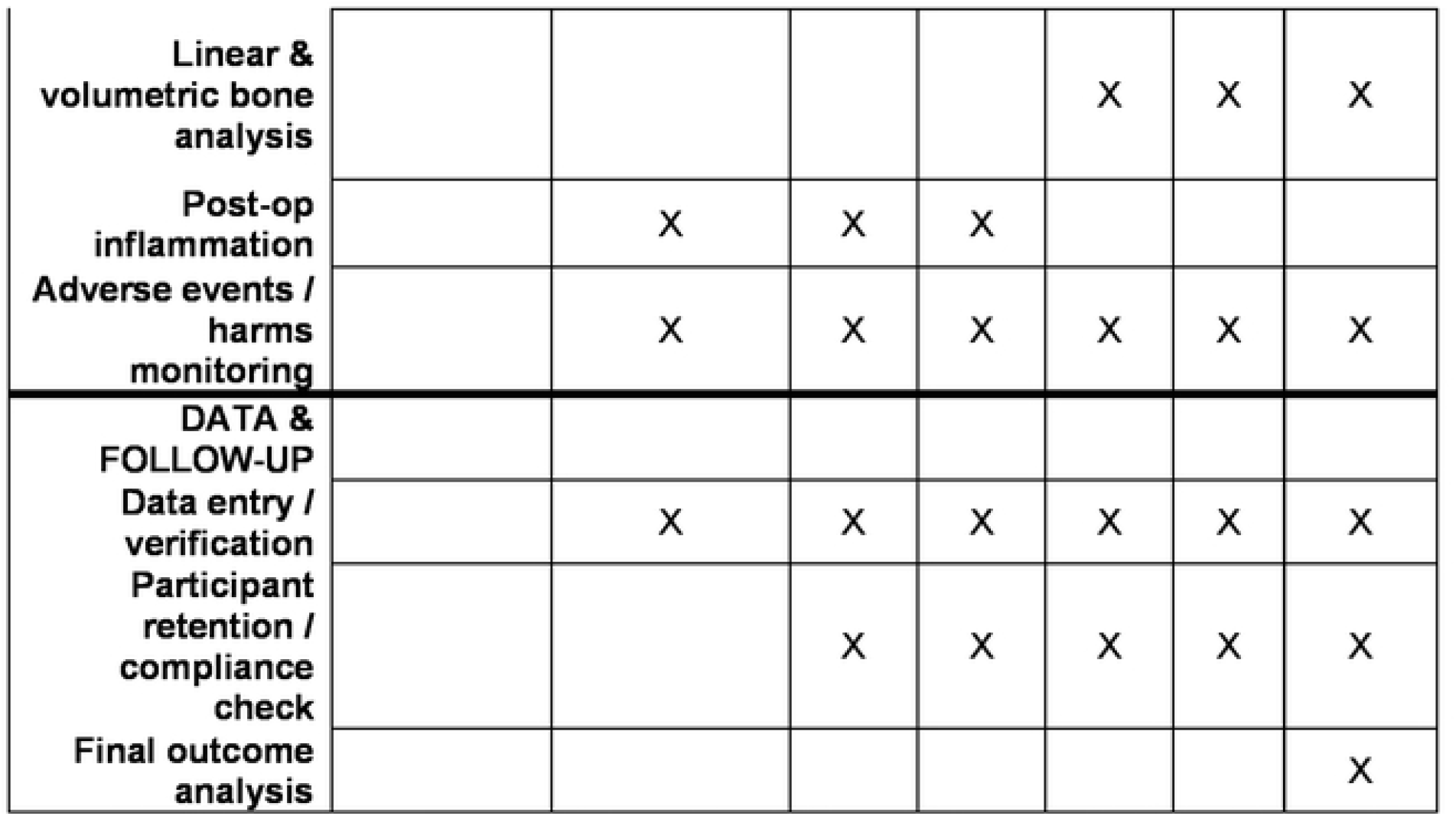
SPIRIT 2025 Schedule of Enrolment, Interventions, and Assessments. Comparison of Bone Augmentation Techniques in the Mandibular Posterior Region: Randomized Clinical Trial Protocol – Schedule of enrolment, interventions, and assessments according to SPIRIT 2025. Legend: t = 0 corresponds to surgical intervention. Follow-up at 1 week, 1, 3, 6, and 12 months. Primary outcome: horizontal bone gain (mm, CBCT). Secondary outcomes: bone density (HU), volumetric ΔVolume, surgical time, postoperative inflammation, and complications.

### Participants

Forty adults (18–64 years) with bilateral posterior mandibular edentulism (Cawood & Howell IV–VI) will be enrolled. All must present good oral hygiene (plaque index < 20%) and systemic health suitable for surgery.

### Exclusion criteria

- Systemic conditions or medications affecting bone healing.
- Smoking > 5 cigarettes/day.
- Pregnancy, lactation, or hormonal contraceptive use.
- Chemotherapy or radiotherapy to the head and neck.
- Active periodontal disease or infection.

Prior to enrollment, all participants will receive both verbal and written explanations of the study objectives, procedures, potential risks, and benefits. Written informed consent will be obtained using a printed physical document signed by hand by each participant before any study-related activity. All signed consent forms will be securely stored in the physical records of the UEES Dental School, accessible only to authorized study personnel.

### Study groups and interventions

Each participant will be assigned to one treatment group only (autogenous, xenogeneic, or allogeneic cortical shell), following simple block randomization in a 1:1:1 ratio.

### Group A – Autogenous cortical shell

Cortical plates (∼1 mm) harvested from the mandibular ramus using a piezoelectric device (*Piezosurgery touch, Mectron, Italy*) will be adapted and fixed with LevelOne® 1.0 mm titanium micro screws (*KLS Martin Group, Tuttlingen, Germany*) made of a Ti-6Al-4V implant-grade alloy (ASTM F136 / DIN ISO 5832-3). These self-retaining Centre-Drive screws are designed for micro-osteosynthesis in oral and maxillofacial surgery, providing excellent torque control and minimal soft-tissue irritation. The enclosed space is filled with a 1:1 mixture of autogenous bone chips and xenogeneic particulate (*Bio-Oss, Geistlich Pharma AG, Switzerland*).

### Group B – Xenogeneic cortical shell

Pre-fabricated bovine laminae (*OsteoBiol® Lamina, Tecnoss®, Italy*) will be rehydrated for five minutes, trimmed to contour, and fixed with screws of the same specifications. The space is filled with the same bone mixture.

### Group C – Allogeneic cortical shell

Human-derived cortical sheets (*Maxgraft® Cortical Sheet, Botiss®, Germany*) will be rehydrated for ten minutes and shaped before fixation with screws. The internal space is filled identically to Groups A and B.

### Surgical procedure

All surgical procedures will be carried out under local anesthesia using Articaine 4% with epinephrine 1:100,000 (*Septodont®, France*). Each intervention will be performed under aseptic conditions with full-thickness flap elevation to expose the posterior mandibular edentulous ridge. A mid-crestal incision will be made along the alveolar crest of the edentulous ridge, followed by two small vertical releasing incisions positioned at the mesial and distal limits of the surgical field to ensure adequate access and flap mobility. The flap will be carefully reflected to expose the bone, avoiding damage to the periosteum at the base of the incision.

The recipient site will be decorticated using a round carbide bur under copious sterile saline irrigation to promote bleeding and facilitate graft revascularization. Small perforations will be made through the cortical plate until medullary bleeding is observed, following the principles described by Khoury and Hanser (2015).

Once the graft corresponding to each experimental group has been positioned and stabilized, and the grafting space filled, a tension-free closure will be performed. The flaps will be passively repositioned after periosteal releasing incisions, ensuring complete coverage of the grafted area and proper adaptation over the augmented ridge.

Wound closure will be achieved using 6-0 blue monofilament nylon sutures (*TechSuture®, São Paulo, Brazil*), a non-absorbable material that provides high tensile strength, precise wound approximation, and minimal tissue reactivity. Interrupted and mattress sutures will be used according to flap configuration to achieve stable primary closure.

Postoperative management will include:

- Antibiotic therapy: *Amoxicillin 875 mg + Clavulanic acid 125 mg*, orally every 12 hours for 7 days.
- Analgesic regimen: *Ibuprofen 600 mg* every 8 hours for 3 days.
- Antiseptic rinse: *0*.*12% chlorhexidine gluconate* twice daily for two weeks.

Patients will be instructed to maintain a soft diet and avoid mechanical trauma in the grafted area. Sutures will be removed 14 days after surgery.

Follow-up visits will be scheduled at 1 week, 1 month, 3 months, 6 months, and 12 months postoperatively. At each follow-up, clinical evaluation of wound healing, tissue integrity, and possible complications (e.g., dehiscence, infection, graft exposure) will be documented. Measured outcomes will include bone gain (mm^3^), bone density (HU), surgical duration (minutes), and postoperative inflammation (mm), the latter assessed as the linear distance from the lip commissure to the tragus on days 3, 7, and 14 after surgery.

### Volumetric and morphometric analysis

Quantitative three-dimensional evaluation will be conducted through a standardized digital workflow that integrates intraoral surface scanning and CBCT-based volumetric analysis. This multimodal approach ensures reproducible evaluation of graft remodeling and contour stability over time.

#### 1. Intraoral scanning

Each participant will undergo digital impression acquisition using a TRIOS® 3 Wired intraoral scanner (*3Shape A/S, Copenhagen, Denmark*) before surgery and at each follow-up interval (3, 6, and 12 months). The scanner provides high-resolution color models (accuracy < 20 μm) that enable precise surface documentation. The resulting STL files will serve to evaluate external volumetric changes and soft-tissue adaptation over time.

#### 2. CBCT-based segmentation and linear measurements

Preoperative and postoperative cone-beam computed tomography (CBCT) scans will be obtained using the Planmeca Viso® G7 CBCT system (*Planmeca Oy, Helsinki, Finland*) under standardized exposure parameters and patient positioning. DICOM datasets will be segmented in the Romexis® 3D module to isolate the grafted region of interest. Surface meshes of the reconstructed bone will be exported as STL files (1:1 scale) for morphometric analysis.

In addition to volumetric evaluation, linear bone measurements will be performed directly in Romexis® following the methodologies described by Monje et al. (2015) and Amaral Valladão et al. (2020).

- For horizontal gain, a perpendicular reference line will be traced relative to the alveolar ridge inclination, and three horizontal lines will be drawn at 5, 7, and 11 mm from the alveolar crest to measure ridge width at each level.
- For vertical gain, measurements will extend from the most coronal point of the alveolar crest to the upper border of the mandibular canal or basal cortical plate, depending on the anatomic limits of each case.
- Each measurement will be performed by two calibrated and blinded examiners, repeated twice with a two-week interval to evaluate intra- and inter-examiner reliability (ICC > 0.9).
- The resulting data will be expressed as the mean horizontal and vertical bone gain (mm) and compared among the experimental groups.

This standardized approach, based on Monje et al. (2015) and Amaral Valladão et al. (2020), allows reproducible quantification of linear bone gain and complements the three-dimensional volumetric workflow performed with Romexis and CloudCompare, ensuring geometric and clinical accuracy in mandibular bone-augmentation assessment.

#### 3. Surface registration and quantitative analysis

STL models obtained from both the TRIOS 3 intraoral scans and Romexis segmentations will be imported into CloudCompare (*EDF R&D, Paris, France*) for alignment and measurement. Surface registration will be performed using the *Iterative Closest Point (ICP)* algorithm (overlap 0.8, max iterations 100, sampling 50 000, RMS < 0.2 mm). Color-coded *Cloud-to-Mesh* distance maps (0–2 mm) will visualize local contour differences and bone volume changes between time points. This enables assessment of both hard-tissue (bone) and soft-tissue remodeling in the grafted regions.

Volume computation will be performed using the *Compute Mesh Volume* function, and the volumetric change (ΔVolume = Vol_post – Vol_pre) will be recorded for each site. All analyses will be repeated twice to ensure measurement reliability, with a mean registration error below 0.1 mm. Quantitative data will be exported for statistical comparison.

This integrated workflow, combining intraoral scanning, CBCT and surface-based morphometric evaluation, provides a transparent, reproducible, and fully digital protocol for monitoring bone and soft-tissue remodeling following cortical shell augmentation.

#### 4. Morphology analysis

Surface-based morphometry will be performed by rigidly aligning pre- and postoperative CBCT-derived meshes on anatomically stable mandibular regions (lingual cortical base and basal mandibular contours) to avoid bias from the augmented site. After alignment, point-to-surface distance maps will be computed to generate color-coded deviation maps (positive values = apposition; negative values = resorption). Morphological metrics will include mean absolute distance (MAD), root-mean-square (RMS) distance, 95th-percentile distance, and the signed volume of positive/negative deviations within a standardized region of interest (ROI) that envelopes the augmented ridge. Mesh preprocessing (isotropic remeshing, light Laplacian smoothing) and ICP registration parameters will be pre-specified. Two blinded examiners will repeat the full pipeline twice (two-week interval); intra-/inter-rater reliability will be reported (ICC).

This surface-based approach complements linear and volumetric outcomes and localizes remodeling patterns across the augmented contour, following validated CBCT subtraction and surface-registration workflows for alveolar augmentation (10,11).

If shape-based correspondence analysis is performed, SPHARM-PDM algorithms may be applied to assess 3D curvature and topographic variations, as described by Kim et al. (12).

### Randomization and blinding

Participants will be allocated to one of the three treatment groups using a pre-specified rotating assignment schedule prepared by an independent coordinator before enrollment, following the sequence A, B, C, and then repeating in the same order. The schedule will be printed as a sequential, sealed list and stored by the study coordinator; investigators will open the next envelope only after confirming eligibility and obtaining consent, thereby preventing foreknowledge of future assignments and minimizing selection bias.

If a participant presents two eligible mandibular posterior sites, the surgical workflow will start on the left side and proceed according to the next assignment in the rotating schedule, while both sites will be managed under the participant’s allocated treatment to preserve the parallel-group design. This standardized left-to-right order is used solely to avoid operational errors and additional bias in timing or side preference.

Outcome assessors (CBCT segmentation, linear measurements, and 3D analyses) and statisticians will remain blinded to group allocation; the surgeon cannot be blinded for technical reasons.

### Sample size

The trial is powered to test non-inferiority of xenogeneic (B) and allogeneic (C) cortical shells versus the autogenous control (A) on horizontal bone gain (mm) at 12 months. Based on CBCT-based randomized clinical trials reporting standard deviations of approximately 1.1–1.4 mm for ridge-width changes following augmentation(13,14), and assuming a common SD of 1.2 mm, a one-sided α = 0.025 and 80 % power, 15 participants per group are required when the non-inferiority margin (Δ) is set at 1.23 mm for a single primary comparison. To preserve family-wise error across the two primary comparisons (B vs A and C vs A), non-inferiority will be assessed in a hierarchical sequence (B vs A first; if met, then C vs A), thus avoiding alpha splitting. The primary analysis will use ANCOVA adjusting for baseline ridge width to reduce variance; under an anticipated baseline–follow-up correlation of ≈ 0.6 (σ_effective ≈ 0.96 mm), a margin of ≈ 1.10–1.20 mm remains adequately powered with n = 15 per group. A blinded, pre-planned variance check will be performed after 20 % enrollment without unblinding group means; if σ_effective exceeds 1.2 mm, a contingency increase to n = 18 per group will be considered.

### Statistical analysis

All statistical analyses will be conducted using IBM SPSS Statistics v28.0 (IBM Corp., USA). Data distribution will be assessed with the Shapiro–Wilk test.

For the primary outcome (horizontal bone gain in mm), a one-way ANOVA will compare mean values among the three groups, followed by Tukey’s post-hoc tests for pairwise differences.

For repeated measurements over time (3, 6, and 12 months), a linear mixed-effects model (LMM) will be applied with Group and Time as fixed effects and Patient as a random effect to account for within-subject correlation. Interaction terms (Group × Time) will be included to evaluate longitudinal trends.

For morphometric outcomes,including mean absolute distance (MAD), root mean square (RMS) deviation, and 95th-percentile distance values obtained from surface-based analysis, group comparisons will be performed using one-way ANOVA (or Kruskal–Wallis for non-normally distributed data). Longitudinal changes in these morphometric parameters will also be analyzed with LMMs incorporating Group, Time, and Group × Time interaction effects.

Categorical variables (e.g., presence of complications) will be analyzed using the Chi-square or Fisher’s exact test, as appropriate. Effect sizes (η^2^ partial for ANOVA, Cohen’s d for pairwise contrasts) and 95% confidence intervals will be reported. A p value < 0.05 will be considered statistically significant.

Missing data exceeding 10% will be handled by multiple imputation under a missing-at-random assumption.

### Ethics approval and monitoring

This study was approved by the Comité de Ética de Investigación en Seres Humanos del Instituto Superior Tecnológico Portoviejo (CEISH-ITSUP), Ecuador (Approval No. 1715824376). All procedures will comply with the Declaration of Helsinki and national regulations. Written informed consent will be obtained from all participants prior to any study procedure. The ethics committee will receive semiannual progress reports; any protocol amendments will be submitted for review and approval before implementation. Adverse events will be recorded and reported according to institutional policy.

### Data management and confidentiality

Clinical and radiographic information will be entered into encrypted electronic case report forms (eCRFs) with double data entry to minimize transcription errors. Each participant will be assigned a unique study code; identifying data will be stored separately from clinical outcomes. De-identified datasets and STL files will be kept on a secure UEES institutional server with access restricted to authorized study staff. After publication, de-identified data will be made available upon reasonable request to the corresponding author, in line with PLOS ONE data policies.

### Expected Results and Discussion

It is anticipated that xenogeneic and allogeneic cortical laminae will achieve bone formation and dimensional stability comparable to autogenous plates, while reducing donor-site morbidity and operative time. Autogenous cortical shells may show faster early integration given their osteogenic potential, yet prefabricated laminae are expected to reach similar volumetric stability under standardized conditions.

The integrated digital workflow is designed to deliver objective, reproducible quantification of remodeling. By combining linear metrics (15,16) with volumetric ΔVolume and colorimetric deviation maps, the study can detect subtle differences across materials beyond conventional 2D measurements.

Limitations include the single-center setting and 12-month follow-up, which may not capture long-term remodeling beyond one year. The parallel-group design ensures independence between interventions and allows direct statistical comparison among three distinct grafting materials under identical surgical and analytical conditions.

## Data Availability

No datasets were generated or analyzed during the current study. Deidentified research data will be made publicly available in the institutional repository of Universidad de Especialidades Espí;ritu Santo (UEES) upon study completion and publication. The study protocol, statistical analysis plan, and informed consent form have been included as Supporting Information files.

## Trial Registration

ClinicalTrials.gov Identifier: NCT06661447

## Acknowledgments

The authors received no funding for this study and declare no competing interests

## Notes

### Competing Interest Statement

The authors declare that they have no competing interests. They have no financial or personal relationships that could inappropriately influence (bias) this work. The study received institutional support from Universidad de Especialidades Espíritu Santo (UEES), which had no role in the study design, data collection and analysis, decision to publish, or preparation of the manuscript.

### Clinical Trial

NCT06661447

### Clinical Protocols

https://clinicaltrials.gov/study/NCT06661447

### Funding Statement

The author(s) received no specific funding for this work.

### Author Declarations

This study was reviewed and approved by the Comité de Ética de InvestigaciÓn en Seres Humanos del Instituto Superior Tecnológico Portoviejo (CEISH-ITSUP), Ecuador (Approval No. 1715824376). The protocol titled “Comparison of Bone Augmentation Techniques in the Mandibular Posterior Region: A Randomized Clinical Trial” was approved for execution at the Dentistry Clinic of the Universidad de Especialidades Espíritu Santo (UEES). All participants will provide written informed consent prior to enrollment, in accordance with the Declaration of Helsinki (2013 revision) and national regulations.

